# Knowledge, Attitudes, and Practices Related to Avian Influenza (H5N1) After the Outbreak in Rural, Cambodia

**DOI:** 10.1101/2023.09.25.23296059

**Authors:** Daraden Vang, Darapheak Chau, Kimim Vutha, Samnang Um

## Abstract

From 2003 to 25 February 2023, the avian influenza (H5N1) virus was confirmed in 59 human infections, including 39 deaths (∼66% case-fatality rate) reported in 13 of 25 provinces in Cambodia. We aimed to assess current knowledge, attitude, and practice toward changes in poultry handling behaviors, poultry consumption, and poultry mortality reporting among rural villagers in areas affected by Avian influenza (H5N1) in Cambodia. A cross-section survey was conducted in August 2023. There were 208 participants residing in Prey Veng province who were invited to be interviewed face-to-face. Descriptive statistics were performed using STATA V17. The participants’ average age was 55 years old (SD=13.3 years), 78.4% were female, 59% had completed primary school, 56.7% were farmers, 68.3% raised chickens in their backyards, and 10.2% raised ducks, 23% of participants cooked sick or dead birds for their families, 32% knowing information about avian influenza virus was a lower proportion from healthcare providers, 10.6% from village health support groups were, and 2% from village animal health workers were only, 49% have been reported poultry illness and deaths to local authorities. The avian influenza epidemic in Cambodia is a genuine threat to animals and a possible concern to humans. To prevent and control this, we strongly advise everyone who works with poultry or wild game birds always to be prepared to follow appropriate hygiene standards and to cook poultry meat properly.

## Introduction

Highly pathogenic avian influenza A (HPAI), also called avian flu or bird flu, is a highly contagious disease of birds caused by avian influenza (AI), a virus of subtype H5N1, that occurred across Asia, Africa, and Europe [1]. It spreads through domesticated and wild birds and is transmitted from poultry to poultry and occasionally from poultry to humans, generally through close contact with infected birds and inhalation of dust or droplets containing the viruses [2].

Globally, from January 2003 to 31 May 2023, 876 cases of human infection with avian influenza A(H5N1) virus were reported from 23 countries [3]. Of these 876 cases, 458 were fatal (CFR of 52%) [3]. Southeast Asia has been identified by the World Health Organization (WHO) as being at risk for emerging diseases, including avian influenza [1,4]. Vietnam has been reported to have the highest number of avian influenza A (H5N1); of these 128 cases, 64 were fatal (CFR of 50%). China has reported the most elevated number of avian influenza A (H5N1); of these 55 cases, 32 were fatal (CFR of 58.2%) [3], while in Cambodia, there were confirmed 58 human infections with the avian influenza A (H5N1) virus have been reported since 2005, including 38 deaths (CFR of 66%) [3]. Several factors contribute to Southeast Asia’s vulnerability to infectious diseases, including population growth and movement, urbanization, environmental factors such as agriculture, land use, water and sanitation, health system factors, and the development of drug resistance [5].

Previous studies have demonstrated that live poultry markets play an essential role in AI transmission from wild birds to humans [6-8]; live poultry markets are high-risk locations for human infection with AI [9]. Therefore, poultry workers, elderly consumers, or those with compromised immune systems visiting the live poultry markets might be at high risk of infection. Despite the utility of preventive behaviors to aid poultry workers and customers in avoiding disease during outbreaks of AI, these preventive behaviors are adopted at low rates by the public, and these low adoption rates are influenced by their perceptions of the effectiveness of control measures [10], infectiousness and severity of the disease [11], and reliability of the information provided by public health authorities [12]. Thus, learning about the public’s knowledge, attitudes, and behaviors is crucial to improving communication efforts by public health officials.

On 23 February 2023, WHO-Cambodia confirmed a new case of human infection with the avian influenza A (H5N1) virus [6]. The case was an 11-year-old girl from Prey Veng province in south Cambodia. No evidence of human-to-human transmission has been reported in Cambodia [6]. Backyard poultry (chickens) were affected at 59 outbreak sites in 14 of 25 provinces reported since 2003 [6]. The response in Cambodia was to cull all poultry, restrict poultry movement and import from other countries, and conduct intensive influenza-like illness surveillance and information, education, and communication (IEC) campaigns within affected provinces [13]. IEC campaigns included leaflets distributed in affected areas and broadcast media coverage on local television and radio. The campaign was designed to inform the public through messages aimed at reducing exposure to disease, preventing the spread of disease in poultry, and encouraging reporting. Additional IEC messages were aired nationally and local news media widely reported outbreaks. Although published reports are available about HPAI-related KAPs, studies have yet to be written on villagers in Cambodia. Therefore, this study assesses current knowledge, attitude, and practice toward changes in poultry handling behaviors, poultry consumption, and poultry mortality reporting among rural villagers in areas affected by Avian influenza (H5N1) in Cambodia. This baseline information about their awareness of HPAI could help policymakers and future researchers with adequate planning and implementation. We assessed current knowledge, attitude, and practice towards changes in poultry handling behaviors, poultry consumption, and poultry mortality reporting among rural villagers in areas affected by Avian influenza (H5N1) in Cambodia.

## Methods

### Ethics Statement

This study was approved by the National Ethical Committee for Health Research, Cambodia. Moreover, this study was keeping participants’ responses strictly confidential. The participants’ names and other personal identifiers are not recorded; privacy is ensured. The written informed consent form was read and given to participants with the overview statement explaining the purpose of the survey.

### Study setting

We conducted a cross-sectional study from 4^th^ to 6^th^ August 2023 at Romleach commune, Sithor Kandal district, Prey Veng province, Cambodia. About 20 km from Prey Veng town, encompassing 382 households and 1,952 inhabitants, 85% of families raise small-scale backyard poultry farming. On 23 February 2023, the Cambodia Disease Control Department notified a new confirmed case of human infection with avian influenza A (H5N1) virus to World Health Organization. A case was an 11-year-old girl residing in Romleach commune, Sithor Kandal district, Prey Veng province, with developed signs and symptoms and received treatment at a local clinic on 16 February 2023. Then, a case was admitted to the National Pediatric Hospital with severe pneumonia on 21 February 2023. A sample was collected the same day through the severe acute respiratory infection (SARI) sentinel system and tested positive for avian influenza A (H5N1) virus by the reverse transcriptase-polymerase chain reaction (RT-PCR) at the National Institute of Public Health on the same day. The patient died on 22 February 2023. Laboratory investigations confirmed the second case on 23 February 2023, the father of the index child. The asymptomatic father is in isolation at the referral hospital [13].

### Study population

We included 208 subjects residing in the outbreak village for at least one year before the survey aged 18 to 79 years who volunteered to provide written or verbal informed consent to participate in the survey interview because they had the opportunity to attend various activities that have been done so far and to obtain information on avian influenza A(H5N1) from it. And who were involved in many households works that are vulnerable or risk them to be in contact with poultry, such as house-keeping, taking care of their children, cooking food for the family members, educating their children, raising poultry, selling poultry, cleaning outdoors, and indoor around their houses.

### Sampling selection

Convenience sampling was used to recruit the participants for face-to-face interviews. After receiving the verbal agreement from the participant, the informed consent was read and provided to the participant. The participants who refused to be interviewed were excluded from the data collection. Only those willing to participate were given informed consent and proceeded to the interview. The interview was performed in a suitable place without any disturbance from outsiders.

### Questionnaire development

The questionnaire is developed with a few modifications of the previously published questionnaire [14-16]. Some new additional questions were invented and created by the local team to fit the local context. The questionnaire was designed in four sections: Section 1 contains demographic information; Section 2 focuses on knowledge of avian influenza A(H5N1), which is composed of 11 questions; Section 3 mentions attitudes related to avian influenza A(H5N1), which contains six questions, and Section 4 describes the prevention practices from avian influenza A(H5N1) which set with 16 questions [17,18].

### Data collection

The eight well-trained public health professionals collected the data through face-to-face interviews using tablets. The duration of each questionnaire was taking approximately 20 minutes to complete the questionnaires. The interview was conducted in Khmer with average speed to reading. During the data collection, participants who agreed to participate were asked to sign an informed consent form.

### Data management

The Khmer-translated questionnaire was stored online using KoboToolbox (https://kf.kobotoolbox.org). KoboToolbox is an open-source software with a free-of-charge server and provides online storage. Interviewers could not access the data once the questionnaires were completed and submitted to the electronic database. Then, the data was exported into STATA version 16 for cleaning and analysis.

### Data analysis

All statistical analyses were performed using STATA version 16 (Stata Corp, College Station, Texas, USA). A descriptive statistic was applied for categorical data as frequency and percentages. For normally distributed data, continuous data is presented as mean and standard deviation (SD).

## Results

### Socio-demographic characteristics of the study samples

A total of 208 subjects from Romleach commune, Sithor Kandal district, Prey Veng province, Cambodia, were included in the analyses. The mean age of participants was 55 years old (SD = 13.3 years), in which age group 21-54 accounted for 45%. The majority, 78.4%, were female. Almost 59% had completed primary education, and 17.3% did not receive formal schooling. 56.7% of the participants were farmers, and 6% were unemployed. Of the total sample, 83.1% of participants had bicycles, 74.4% had a smartphone, 73.9% had motorbikes, and 53.6% had television. Almost 68.3% raise chickens’s backyard, and 10.2% raise ducks (see **Table 1**).

**Table 1.** Socio-demographic characteristics of subjects (n=208)

| Characteristics | Frequency | Percent |
| --- | --- | --- |
| **Gender** |  |  |
| Male | 45 | 21.6 |
| Female | 163 | 78.4 |
| **Mean age in years (SD)** | 55(13.3) |  |
| 21-39 | 27 | 13.0 |
| 40-54 | 66 | 31.7 |
| 55-64 | 65 | 31.3 |
| 65-79 | 50 | 24.0 |
| **Education** |  |  |
| None | 36 | 17.3 |
| Pagoda/Primary | 122 | 58.7 |
| Secondary | 37 | 17.8 |
| Higher and above | 13 | 6.3 |
| **Occupation** |  |  |
| Farmer | 118 | **56.7** |
| Housewife | 39 | 18.8 |
| Self-employed/home business | 30 | 14.4 |
| Unemployed | 12 | 5.8 |
| Teacher/Healthcare provider | 9 | 4.3 |
| **Asset Ownership** |  |  |
| Bicycle | 172 | 83.1 |
| Smart Phone | 154 | **74.4** |
| Motorbike | 153 | 73.9 |
| TV | 111 | 53.6 |
| Radio | 70 | 33.8 |
| Car | 11 | 5.3 |
| **Poultry Ownership** |  |  |
| None | 60 | 29.3 |
| Chickens | 140 | **68.3** |
| Ducks | 21 | 10.2 |
| Any poultry | 1 | 0.5 |

### Self-reported practices in poultry illness or died

More proportions (97.1%) of adults reported burying dead poultry; 23% said they prepared food for the family, 8.2% threw dead poultry into water sources, and only two percent reported burning dead poultry (see **Table 2**).

**Table 2.**
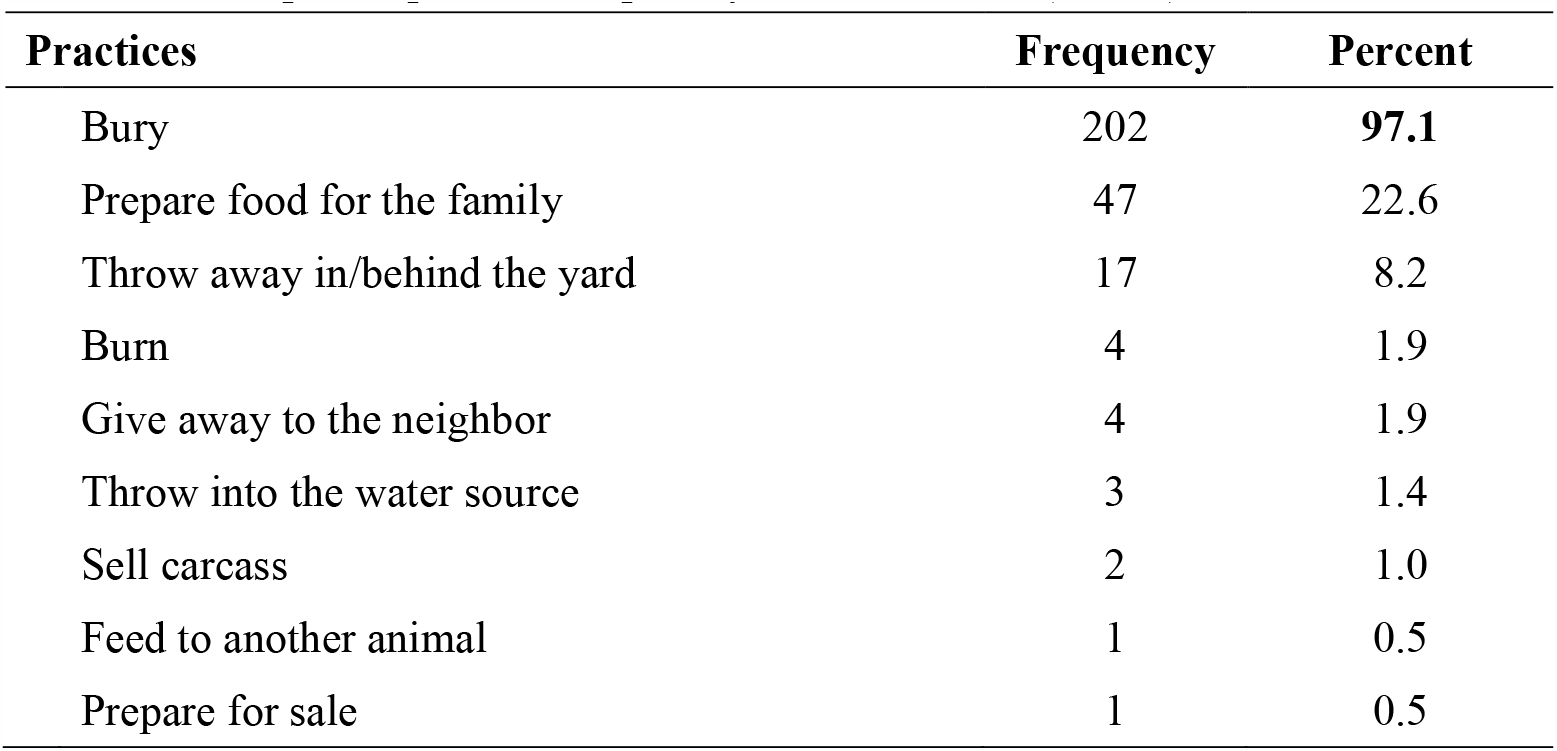
Self-reported practices in poultry illness or death (n=208)

### Source of avian influenza information

Illustrated **Fig**. 1 shows the self-reported multiple sources of avian influenza information. A friend or relative was the first source that could provide knowledge on avian influenza for most people (39%), followed by television (36%), the village chief and healthcare provider (32%), and social media (23%).

**Fig 1.**
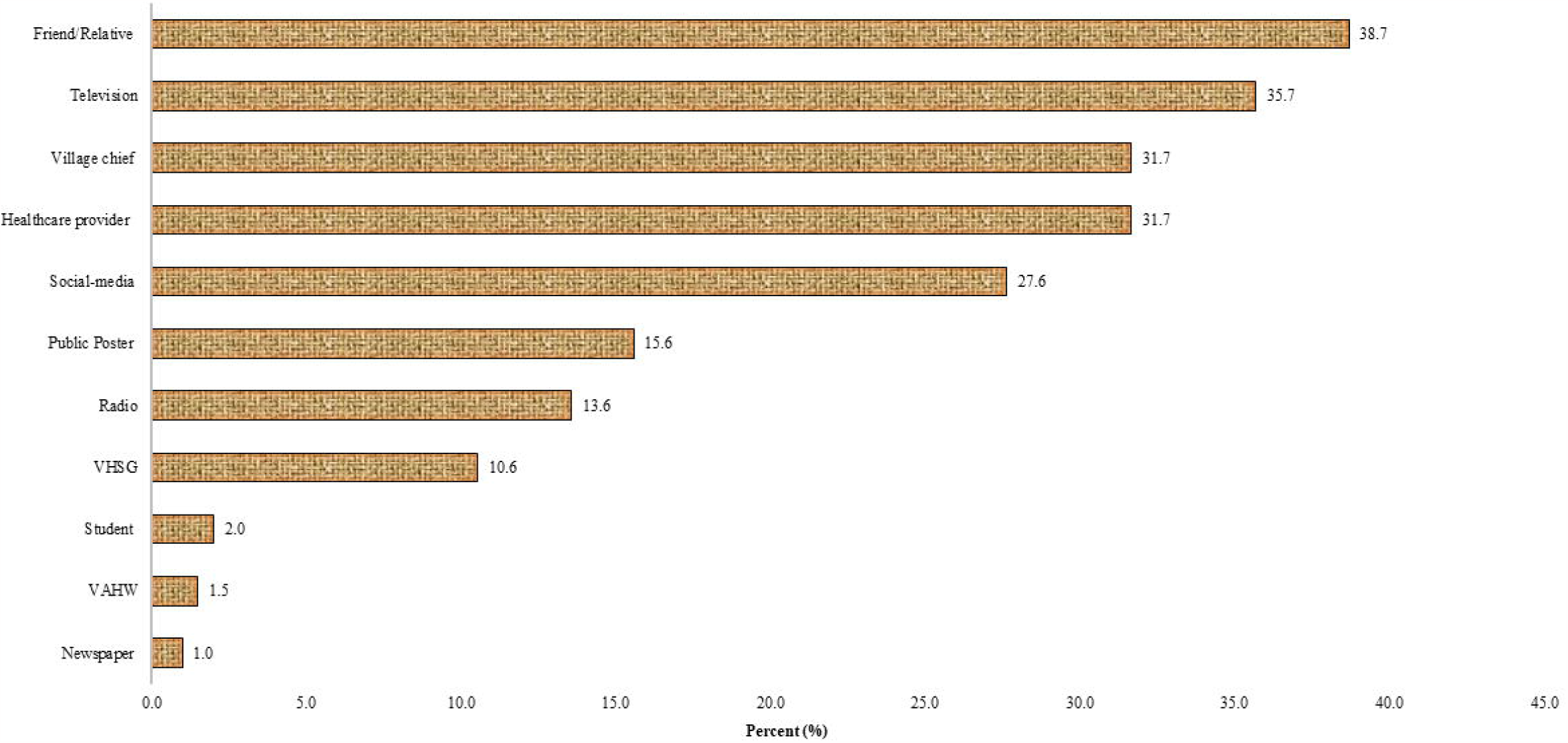
Self-reported multiple sources of avian influenza information (n=199); **\***VHSG=Village Health Support Groups; *VAHW=Village Animal Health Workers.

### Knowledge of sick/dead poultry reported

Fewer individuals reported sick or dead poultry in their backyards. 49% of participants reported ill or dead poultry to village leaders, whereas 47% did not (see **Fig 2**).

**Fig 2.**
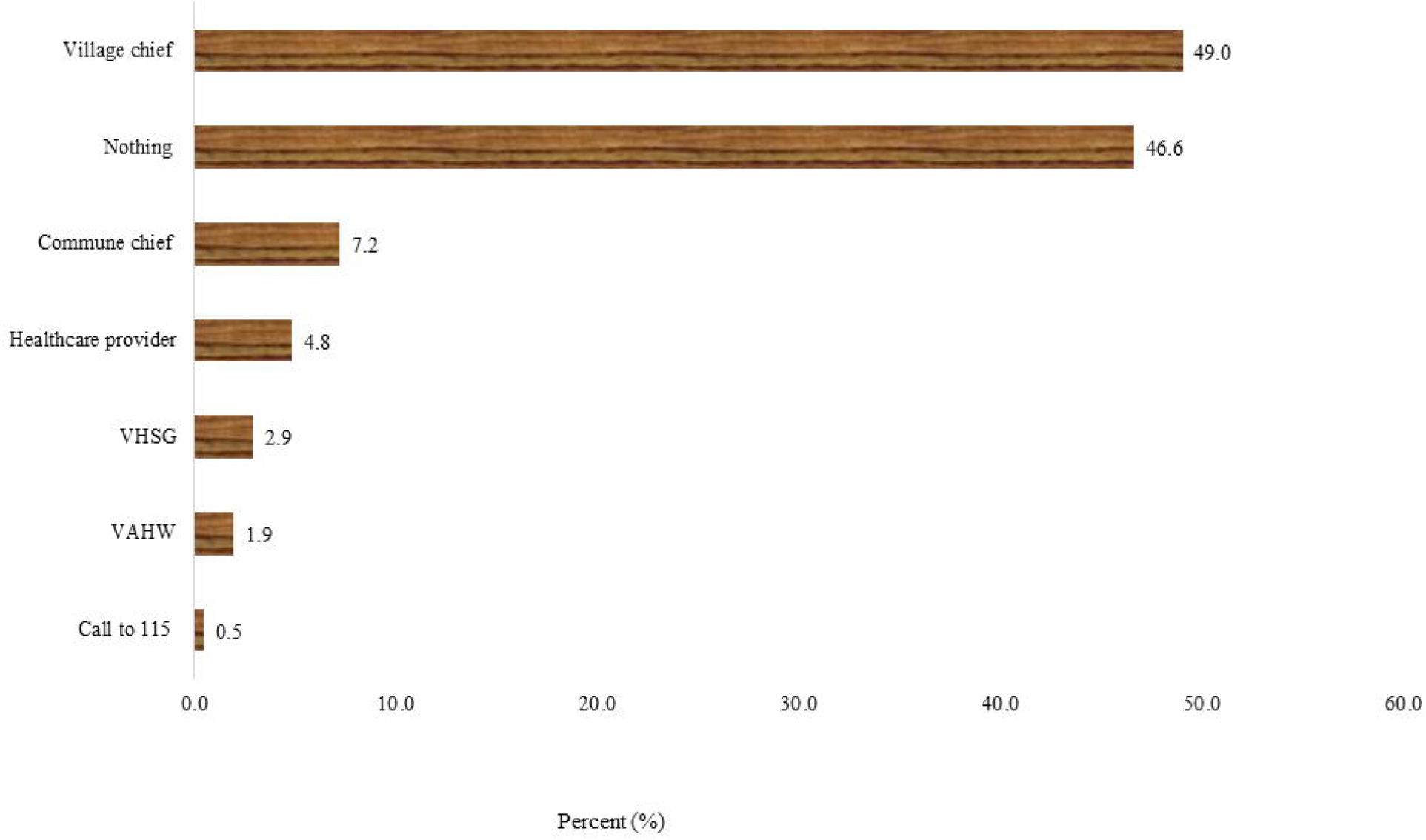
Self-reported multiple sources of reported sick/dead poultries (n=208); **\***VHSG=Village Health Support Groups; *VAHW=Village Animal Health Workers.

The proportion of people who reported interacting with poultry. 20.4% provided care or assistance to poultry, 26.6% purchased poultry from the market for food throughout the study period, and 41.1% used poultry excrement for manure (see **Table 3)**.

**Table 3.** Self-reported with poultry contact among participants (n=207)

| **Reported Practice** | **Frequency** | **Percent** |
| --- | --- | --- |
| **Contact with domestic poultry** |  |  |
| Care or help care for poultry | 42 | **20.4** |
| Touch sick or dead poultry with bare hands | 19 | 9.2 |
| Allow children in the household to play (touch and catch) with poultry | 8 | 3.9 |
| Slaughter poultry | 5 | 2.4 |
| Use dead domestic poultry from the yard for household consumption | 4 | 1.9 |
| **Contact with poultry at live bird markets** |  |  |
| Ever bought poultry from the market for food during the study period | 55 | **26.6** |
| **Contact with wild birds** |  |  |
| Eat wild birds | 3 | 1.5 |
| Collect dead wild birds from the field for household consumption | 2 | 1.0 |
| Ever prepared wild birds for food | 2 | 1.0 |
| **Potential environmental contamination** |  |  |
| Use poultry feces for manure | 85 | **41.1** |
| Prepare poultry near a pond, river, or water well | 7 | 3.5 |
| Wash poultry products directly in the water source (pond/river) | 7 | 3.5 |

## Discussion

We conducted a cross-sectional survey of 208 subjects in Prey Veng province from 4 August to 7 2023 to assess current knowledge, attitude, and practice regarding changes in poultry handling behaviors, poultry consumption, and poultry mortality reporting. Since 23 February 2023, Cambodia’s Ministry of Health has reported to WHO one confirmed fatal case of human infection with the avian influenza A(H5N1) virus in an 11-year-old girl from Prey Veng province. Then, on 23 February 2023, epidemiological and analytical studies revealed the second case, the father of the index child [13]. Within the affected village, information, education, and communication (IEC) campaigns were implemented, including leaflets distributed and broadcast media coverage on local television and radio to inform the public through messages aimed at reducing exposure to disease, preventing disease spread in poultry, and encouraging reporting.

In our survey, the proportion of subjects who work as farmers was 57%, raising chickens in the backyard was 68.3%, and raising ducks was 10.2%. Cambodia’s higher-risk behaviors or vulnerability groups need priority intervention to reduce infectious and zoonotic diseases. Since the first outbreaks of highly pathogenic avian influenza caused by viruses of the H5N1 subtype (H5N1 HPAI) were detected in geese in 1996 in the Chinese province of Guangdong and 1997 in Hong Kong, Until today, 54 countries in Asia, Europe, and Africa have reported H5N1 HPAI outbreaks among poultry; outbreaks in poultry farms have been reported in every country in Southeast Asia (SEA) [2,6,19,20]. Furthermore, we noticed that 23% of participants cooked sick or dead poultries for their families. Previous research in Cambodia found that direct contact with chickens suspected of being infected with the H5N1 virus was harmful [20]. Of the participants who reported knowing information about the avian influenza virus, a lower proportion from healthcare providers was 32%; village health support groups were 10.6%; village animal health workers were only 2%. Implementing information, education, and communication (IEC) campaigns should be conducting refresher training for villagers. This study found that 49% reported poultry illness and deaths to local authorities. These reported practices did not improve during the study period. Raising backyard poultry in rural Cambodia provides significant income and nutrition with an excellent annual investment. Government recommendations to reduce the risk of avian influenza transmission did not impact the behavior of poultry producers.

Further research should prioritize developing interventions that simultaneously reduce the risk of avian influenza transmission and increase the productivity of backyard poultry [21]. However, closer relationships between local healthcare providers and residents could promote early healthcare behaviors in people living in rural communities. Educational programs conducted by local healthcare providers might be effective. Still, residents’ attitudes must be considered when planning health education in communities with H5N1 patients in neighboring areas [22].

### Limitations

This study was limited to comparing participants living in one community affected and the other unaffected by the avian influenza (H5N1) outbreak in February 2023, representing a rural setting in a Prey Veng province of Cambodia. Participants who did not have household poultry were included in the study participants and they need to answer some questions as if they have household poultry. Further investigations comparing subjects with similar socioeconomic conditions and common educational and environmental backgrounds are required to assess the influence of an H5N1 outbreak further.

## Conclusion

The avian influenza epidemic in Cambodia is a genuine threat to animals and a possible concern to humans. Our findings suggest that a high prevalence (23%) of participants reported cooked sick or dead poultries for their families. A lower proportion of them said that knowing information about the avian influenza virus by healthcare providers was 32%; village health support groups were 10.6%; village animal health workers were only 2%. Similarly, villagers were lower reported (49%) their poultry illnesses and deaths to local authorities. Thus, to prevent and control this, we strongly advise everyone who works with poultry or wild game birds to always be prepared to follow appropriate hygiene standards and to cook poultry meat properly.

## Data Availability

All relevant data are within the paper.

## Acknowledgments

The authors would like to thank the village chiefs and study subjects for participating in our studies and staff from the National Institute of Public Health, namely Mr. **Clo Yong**, Mr. **Pao Pich**, Mr. **Chy Bunthoeun**, Mr. **Pann Meng Heng** for assisting with the survey.

